# Value Set Hub: Software for developing and curating high-quality value sets

**DOI:** 10.1101/2024.10.07.24315009

**Authors:** Sigfried Gold, Joseph E. Flack, Wayne G. Lutters, Christopher G. Chute

## Abstract

Terminology value sets are central to research studies using real-world patient data. We describe the considerable challenges that arise when researchers develop value sets for their studies or reuse those made by others. We present Value Set Hub (VS-Hub), software to overcome these challenges, describing its design, implementation, features, use in the field, lessons learned, and future directions. Over a five-month period, VS-Hub has been used by over 200 users and has been used in the development and curation of 95 recommended value sets for commonly studied conditions, treatments, and lab tests. Particular innovations include the presentation of multiple value sets on the same screen for easy comparison, the display of compared value sets in the context of vocabulary hierarchies, the integration of these analytic features and value set authoring, and value set browsing features that encourage users to review existing value sets that may be relevant to their needs.

## Introduction

The importance and productivity of observational, *in silico* research based on electronic health records, reimbursement claims, and other real-world data (RWD) has exploded over the past ten to fifteen years. Thousands of researchers in networks like OHDSI, PCORNet, All of Us, and N3C^*^ are able to leverage vast, multi-site data sources harmonized to common data models (CDM) using open-source software and infrastructure to perform replicable, FAIR research at hitherto unheard-of speed and low cost.

A particular problem area in the execution of RWD studies is in the development of analytic value sets: groups of controlled medical terminology codes used to query patient records in the computation of cohort or phenotype membership and study variables.^6†^ Designing the algorithms used in executing research or safety studies (to compare outcomes for alternative treatments, for example) requires an understanding of observational, retrospective study design; the clinical topic of interest; and the care settings and operational workflows shaping the data available for study. Formulating the conditional and temporal logic for the overall study and specific cohorts and variables entails unavoidable thought and work. The selection of codes for the value sets used in these algorithms seldom receives as much attention despite the fact that these value sets determine the selection of patient data that serve as input to the algorithms.

Creating value sets can be easy, for instance, by doing a string search for vocabulary terms and then, perhaps, navigating around vocabulary hierarchies to find relevant related terms. This might result in a perfectly adequate value set, or not. It is often not feasible to perform a thorough empirical validation based on a gold standard of patient records marked as having or not having some particular condition or phenotype. Lacking that, the quality of a value set is not directly measurable; it is indirectly inferred by diligently applying the best practices in its creation: thorough reviews by clinical and terminology experts, cross-referencing with similar value sets, review of matching record counts, and spot checking of those records.^7^

As a field, we know these best practices and see occasional papers exhorting us to use them or offering improvements to one or another, yet quality problems persist.^8–10^ Particular attention has been given to the sharing and reuse of value sets^11^ in public value set repositories like VSAC^12^ and Clinical Codes^13^ or repositories integrated into larger RWD research platforms like OHDSI/ATLAS^2^ and the N3C Enclave.^5^

Recognizing the effort and skill required to craft high-quality value sets, the repository developers offer tools to encourage sharing and reuse, hoping that researchers will take advantage of others’ work, building on it where possible instead of repeating it. What we see in actual repositories, however, is a proliferation of value sets for common features of interest (diagnoses, treatments, etc.)^6^ Those needing value sets either do not think to check for existing value sets and create their own or, finding many candidates for reuse with no easy way to discern their quality of appropriateness for their needs, they again make their own.

In the N3C community, we and our colleagues have made concerted efforts to encourage reuse by asking value set authors to provide metadata about the provenance, intentions, and limitations of their value sets and by providing extensive documentation and training to promote best practices and reuse. These efforts have not led to discernable improvement. Because current tools can make it time-consuming and difficult to follow best practices — e.g., expert review of large value sets when concepts are not presented hierarchically; lack of effective interfaces for comparing candidates for reuse; lack of or inconvenient access to term usage counts — much of the work required to make a high-quality value set may not happen.

We have come to believe that the only way to get users to reuse appropriate value sets or, when creating their own, to follow best practices, dedicating sufficient and fitting effort to create them well and prepare them for reuse by others, is to provide software that pushes them to do these things, mostly by making it easy and obviously beneficial to do them.

We present Value Set Hub (VS-Hub)^*^ as a platform for browsing, comparing, analyzing, and authoring value sets — a tool in which the presence of multiple, sometimes redundant, value sets for the same condition strengthens rather than stymies efforts to build on the work of prior value set developers. VS-Hub introduces several innovations to the state of the art for value set authoring platforms.

In the Design section below, we describe the goals and requirements that have driven VS-Hub design and the tools used to build it. The implementation section provides an abbreviated account of the development trajectory, the evolving needs and priorities that have driven implementation of specific features, a sense of the overall architecture, and description of the features on the two primary user interface (UI) screens. The evaluation section gives quantitative and narrative description of VS-Hub’s actual use in the development and maintenance of value sets over the past several months. The Discussion section addresses generalizability and opportunities for further work.

## Design

VS-Hub’s developers work as part of a team whose mandate includes the curation, development, and maintenance of recommended value sets for conditions and electronic phenotypes (i.e., cohort selection or research variable algorithms) commonly needed for RWD research. Understanding that each research project is unique and that researchers will sometimes require more than a one-size-fits-all value set, we have developed VS-Hub both to serve our own office (and other informaticists with terminology expertise who similarly endeavor to build or curate value sets for use beyond a specific project) and to serve the more general audience of those looking to find or create value sets for a specific need.

Specifically, the software should:

- Maximize the information immediately visible or rapidly available to support user decision-making as they review existing value sets and reuse or revise them for their own studies;
- Encourage the user to find and review existing value sets most relevant to their topic before creating a new one, showing them summary data regarding value sets of possible interest; counts of definition and expansion concepts; matching patient and record counts; author, version, intention, provenance, and other metadata;
- Make users aware of the semantic neighborhood of the concepts they are considering by showing (visualizing) value set member concepts in the context of their vocabulary hierarchies and other semantic information when available (e.g., concept domains, mappings, membership in other value sets);
- Provide any available empirical information about individual concepts, such as patient, record, and descendant record counts or validation data;
- Present many-to-many comparison of selected value sets;
- Highlight differences between selected value sets;
- Encourage the development of parsimonious value sets — that is, value sets defined intensionally using a minimal set of high-level concepts that will be expanded to include or exclude their descendants.
- Encourage thoughtful review and maintenance of value sets after vocabulary updates, showing concepts added or removed when expanding definition (intensional) concepts using current vocabulary versions;
- Effectively hide data when value sets include too many concepts for performant display in browsers or comprehension by users (e.g., by collapsing very deep or very wide descendant trees), providing clear summary of hidden information to facilitate discovery and display.

The long-term aim of VS-Hub is to serve as a central value set exchange and authoring platform, interoperable and synchronized with external sources of value sets such as VSAC, N3C, ATLAS instances, and FHIR value set resources. Though it is designed for generalizability, implemented features so far have been tailored to the evolving needs of its initial audience, the thousands of researchers in the N3C community. This community conducts research using the N3C Enclave, a secure environment built and hosted with Palantir Foundry for managing and analyzing harmonized, multisite data for 22 million COVID patients and controls. Its data structures follow the OMOP CDM and it uses the OMOP vocabulary system to harmonize and integrate concepts from many source vocabularies (e.g., SNOMED, RxNorm, LOINC, CPT, ICD10CM, etc.)

After extensive work with Palantir engineers building the Enclave’s Concept Set Browser and Editor, it became clear that many of the aims listed above would not be effectively implemented because 1) engineer time for the project was limited, and 2) the Palantir Foundry UI development tools were not designed for the kind of information-dense display and rapid interactivity we believed necessary. Hence, we determined to build VS-Hub outside the Enclave using standard web development tools. Additional motivations for that choice include being able: to (eventually) accommodate and translate between many value set repositories and frameworks; to allow value set review by subject matter experts who don’t have Enclave accounts and generally serve the wider informatics and research communities; to facilitate rapid feature development using our tools of choice; and to allow and invite open-source contributions from informaticists and software engineers beyond our team.

Software tools and platforms used in the implementation of VS-Hub are listed in Table 1.

**Table 1.** Submission type, abstract length, and page length maximum for AMIA submissions.

| Tool | Type | Purpose |
| --- | --- | --- |
| PostgreSQL | Database server | Backend data management |
| Python | Language | Backend server, external system synchronization, unit testing |
| FastAPI | Python package | Backend server |
| OAK* | Python package | Semantic graph query for vocabulary data |
| NetworkX | Python package | Semantic graph query for vocabulary data |
| JavaScript | Language | Frontend web interface |
| React | JavaScript package | Frontend user interface |
| Graphology | JavaScript package | Semantic graph query for vocabulary and value set data |
| Jest | JavaScript package | Frontend unit testing |
| Playwright | JavaScript package | Frontend end-to-end testing |
| Azure | App hosting service | Hosting of database and backend and frontend applications |
| GitHub | Code hosting service | Version control; management of Azure deployment and continuous integration testing |

\* Ontology Access Kit is cited in the references.<sup>14</sup> The other tools listed are easily found using any search engine.

**Table 1.** Usage and application statistics.

| Measure | Value | Notes |
| --- | --- | --- |
| Total log records | 28,703 | All API calls that invoke tracking |
| Log records -- developer IPs removed | 23,704 |  |
| Log sessions | 12,014 | Calls to multiple API endpoints from the same browser page are grouped together as "log sessions" |
| IP addresses | 253 |  |
| User API call errors | 11 |  |
| Developer API call errors | 183 |  |

## Implementation

Though the bulleted aims listed above have all been achieved to some degree, we have had and continue to have a long list of planned features to implement them more effectively. Design and development have often been driven by specific projects our team has been responsible for and features have been implemented as resources allow. We will give a brief account of the development trajectory.

Our first attempt to provide the community with a library of N3C-recommended value sets was based on importing credible value sets from VSAC. Toward that end, we built a framework for storing value sets, using VSAC APIs for import and Palantir Foundry APIs for upload to the Enclave. This strategy also required conversion from source vocabulary codes to OMOP standard concept IDs. This was straightforward for conversion from vocabularies that OMOP counts as standard (SNOMED, RxNorm, etc.); but from other vocabularies (like ICD10), mapping to standard proved problematic, especially due to pre/post coordination issues.^15^

As these and other issues affected the majority of the value sets we needed, we turned to using and improving value sets already in the Enclave. There we faced a problem of massive redundancy and clutter. For instance, of the 5,000 value sets in the Enclave (7,600 if counting versions), 80 contain the word ‘diabetes’ (260 if counting versions.)

We extended our framework to download and update value sets from N3C and we built a user interface for browsing and selecting from these (including display of relevant metadata), recommending related value sets, comparing those selected to each other, and presenting member concepts with an indented tree based on vocabulary hierarchies. We have struggled and tried many different approaches to dealing with the problems of recommending related value sets, displaying several value sets at a time, and retrieving and displaying the amounts of data involved when handling larger value sets.

**VS-Hub’s search, browse, recommend, and select screen (Figure 1)** treats every selected value set equally, so the related value table presents a list of every value set sharing one or more concepts with any of the value sets selected so far. With the union of all the concepts belonging to the selected value sets as a basis, the related list shows precision and recall figures and other counts, allowing the user to sort on these columns to find related value sets most relevant to their needs. In order to calculate the shared concepts, precision, and recall columns for the three value sets selected in screenshot in Figure 1, we take their total 1,074 distinct member concepts, retrieve the 500 related value sets that contain at least one of those, then retrieve the members of each related value set: 1,225,496 total, 304,472 distinct concepts.

**Figure 1.**
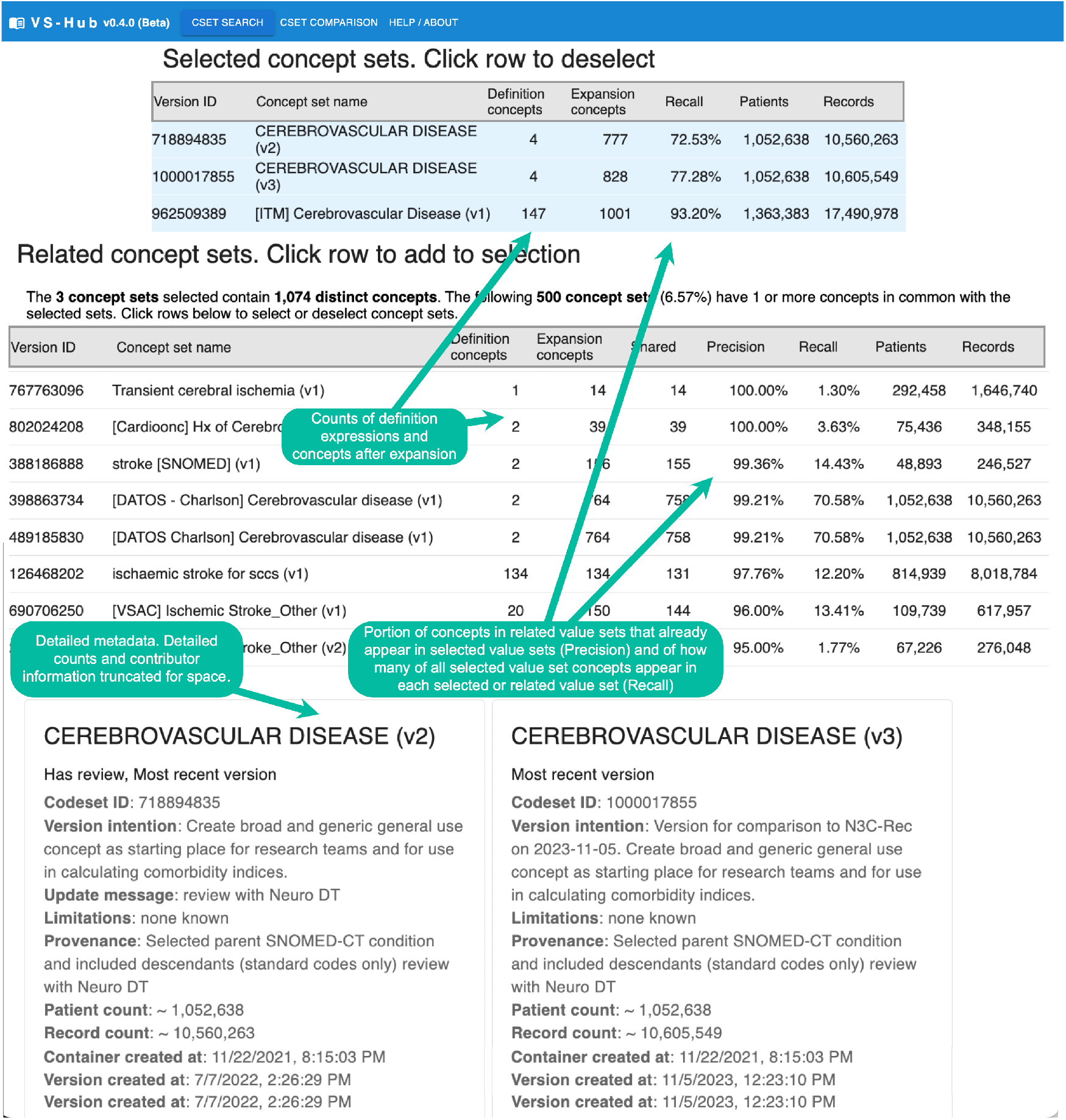
VS-Hub’s search, browse, recommend, select screen.

We give these numbers not to be tedious but to convey that even for moderately sized value sets (100 – 1,000 concepts), the calculation of these figures can involve substantial data processing. In order to keep the application from being painfully slow, we have tried a variety of optimization and caching strategies (discussed below in Lessons learned.)

**VS-Hub’s display, comparison, and authoring page (Figure 2)** presents a table of concepts (first column of each row), metadata (middle columns), and value set membership (rightmost columns.) Another column on the right would appear for constructing a new value set. We omitted that and description of its UI features for reasons of space.

**Figure 2.**
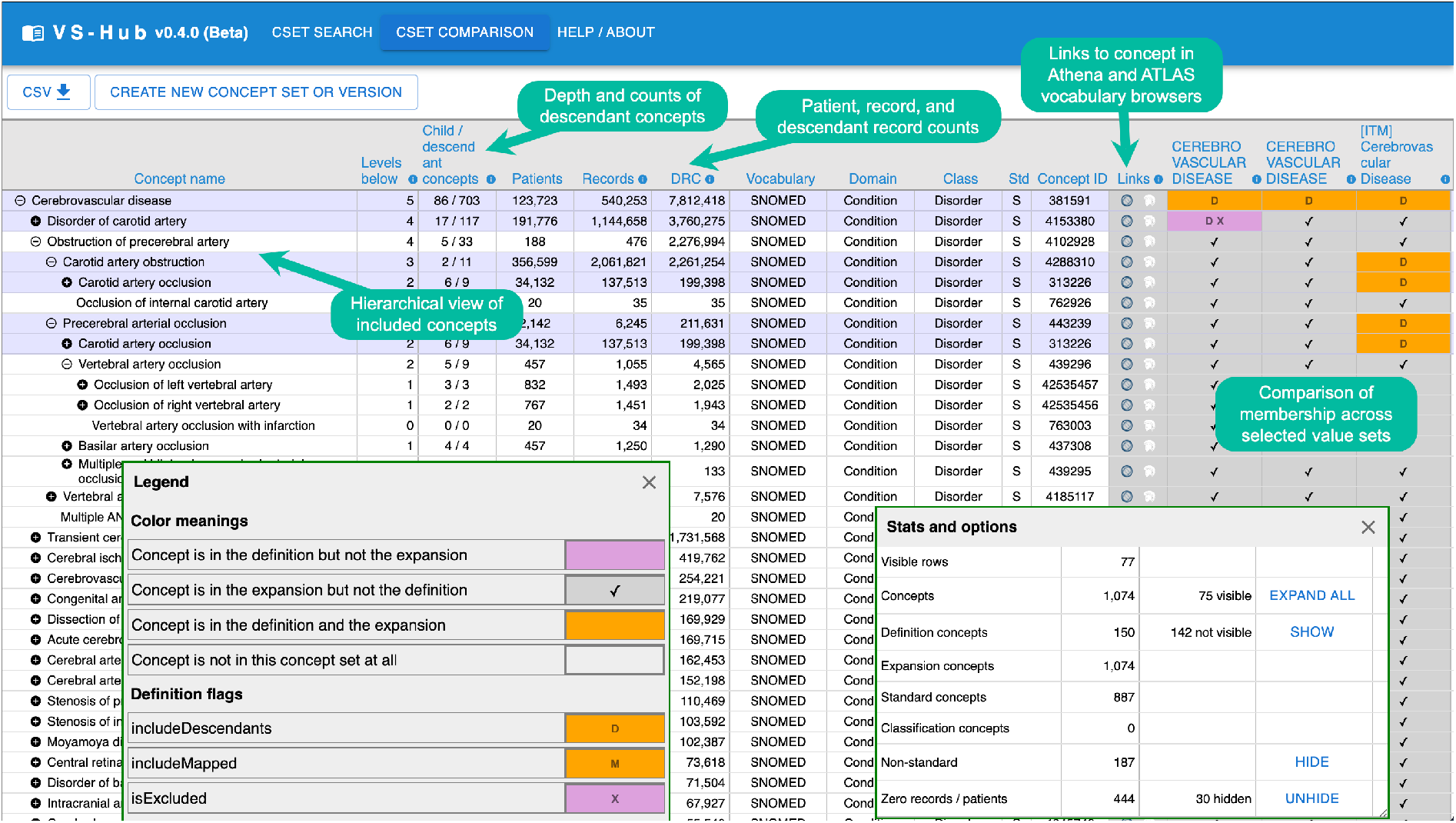
VS-Hub’s display, comparison, and authoring page.

The OMOP vocabulary system used by N3C and many of its source vocabularies are structured as polyhierarchies or directed acyclic graphs (DAG), that is, pairs of concepts (terms, codes) are connected by directed edges, relating them as parent/child or source/target, such that a parent generally has many children and a child sometimes has more than one parent.^*^ Intuitively users think of groups of related concepts as forming a tree, like a file system directory tree. VS-Hub follows the convention of representing such trees as collapsible, nested (indented) lists, though we have had to re-implement the indented list to deal with several complexities.^†^

We use available React components for frontend UI elements where possible. Available nested list controls only allow individual items to be displayed as a single string or React component. Given the amount of information we want to convey about each concept, a tabular display was necessary. We represent nesting by indenting the first column (concept name) of each row and showing an expand/collapse (+/–) icon on rows with children. One shortcoming of our current implementation using react-data-table is that we are only able to add or remove rows by re-rendering the whole table. It would be better to animate the expand/collapse operations.

In order to construct the nested tree display, we select a subgraph of the full OMOP vocabulary DAG^*^ consisting of all the concepts included in the selected value sets and recurse through it, showing concepts indented by level, multiple times if they have multiple parents, and sorting each level by descendant record count or other columns of the user’s choice. Because some value sets are so large that displaying all their concepts will crash the browser tab, the hierarchy is initially displayed with all nestings collapsed, allowing the user to expand individual rows or click Expand All from the Stats and options dialogue. As seen in Figure 2, concepts that appear in the definition of at least one of the selected value sets are shaded in light purple. As indicated in the Legend, concepts that appear in the expansion but not the definition of a value set display a checkmark at the intersection of the concept and that value set. Intensional definition concepts show the options defined for their expansion (D for including descendants, M for including mapped concepts, X for excluding from the expansion — see cells in top right of Figure 2) and are shaded orange or purple depending on whether they appear in the expansion.

A problem we have had to address in the hierarchical display is that sometimes the set of concepts will include pairs that do have an ancestor-descendant relationship but not the intervening concepts that connect them. In that case, the generated subgraph will not contain an edge connecting them and the descendant will act as a root of a disconnected component. Since users want to be aware of relevant ancestor-descendant relationships, we fill in the missing nodes. Getting this right was a trial involving many (mostly misleading) conversations with ChatGPT. Eventually, with much care, we developed a unit test that handles the various edge cases we found in different value sets, diagrammed in Figure 3. The final algorithm works by traversing the full graph upward from each subgraph leaf node, capturing ancestor nodes up to any whole graph root, and discarding any of these that do not appear between two subgraph nodes.

**Figure 3.**
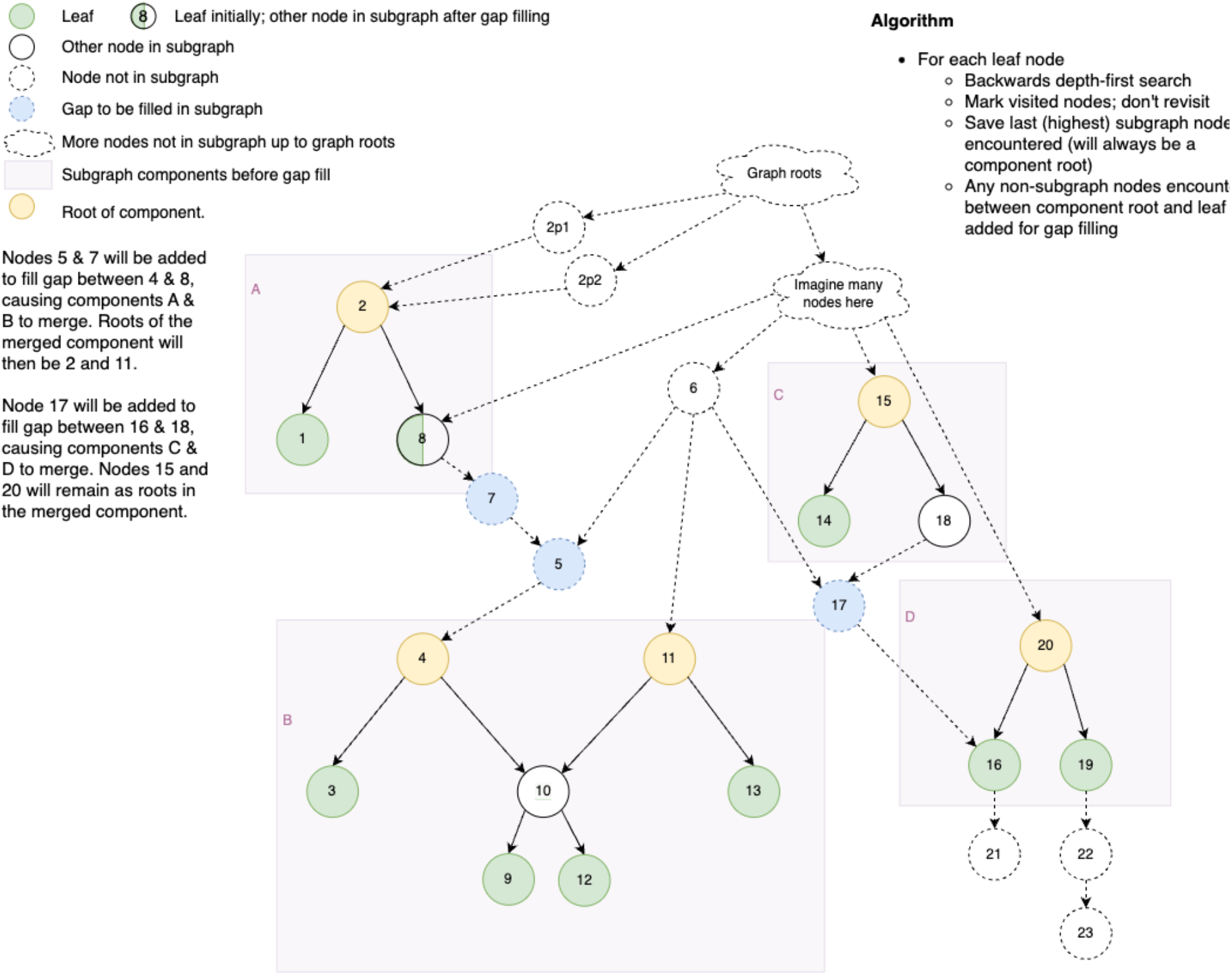
Diagram of gap-filling algorithm.

## Evaluation

Between November 2023 and March 2024, we collected backend server usage logs. Since we use caching to avoid redundant server calls, these logs do not capture analysis of already-downloaded data. After removing 4,999 log entries of testing or use by VS-Hub developers, the remaining 23,704 records represent use by our target audiences. Testing was agile and incremental amidst pilot deployment as beta software. Some of the 23,704 records analyzed include users just trying the software out rather than performing a specific task.) Table 1 provides summary data captured by usage logging. These figures make clear that VS-Hub is being used beyond the team that is building it.

Figure 4 shows a multi-modal distribution of VS-Hub usage levels; that is, over the five months of log capture, about 80 users made fewer than five page visits; about 70 made a few more visits; about 50 made between 15 and 20, and 10 or so made around 40 visits. (This chart does not include use by developers.)

**Figure 4.**
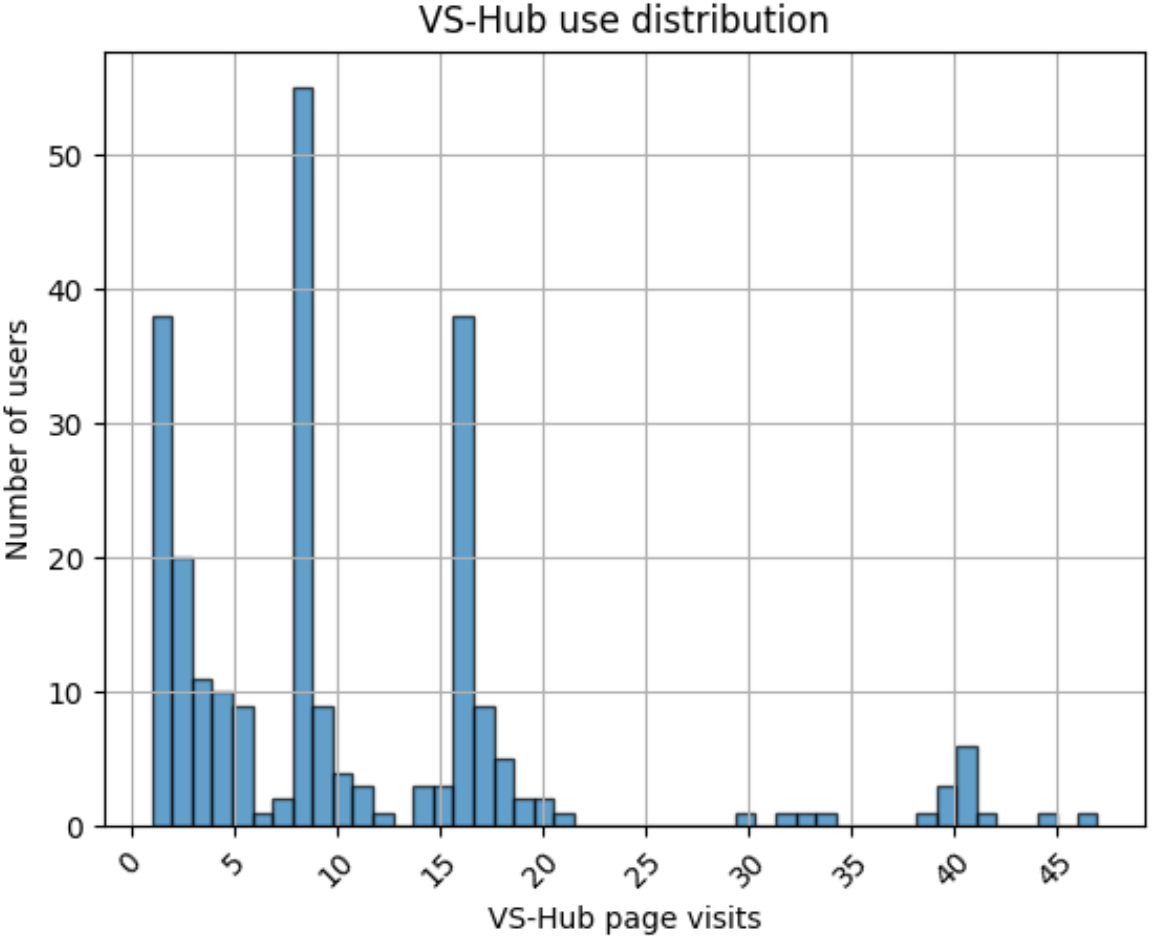
VS-Hub use distribution across users.

Table 2 shows the range of value set sizes in the N3C repository as a whole and the count of concepts (from one or more value sets) returned from VS-Hub API calls that return concepts.

**Table 2.** Concepts in value sets and VS-Hub calls.

| Value set size across N3C |  | Concept counts in VS-Hub calls |  |
| --- | --- | --- | --- |
| Concepts | Value sets | Concepts | VS-Hub calls |
| 1 - 9 | 774 | 1 - 9 | 15 |
| 10 - 99 | 1,862 | 10 - 99 | 1,326 |
| 100 - 999 | 2,596 | 100 - 999 | 620 |
| 1,000 - 9,999 | 1,357 | 1,000 - 9,999 | 129 |
| 10,000 - 99,999 | 537 | 10,000 - 99,999 | 50 |
| 100,000 - 999,999 | 64 | 100,000 - 999,999 | 24 |

## Discussion

### Lessons learned

The hardest problem we faced and where we made the most mistakes was in generating the indented tree display for large value sets. We should have addressed it from the first rather than waiting until code had already been written and users started needing to work with value sets larger than the application could handle. Even then, our initial approaches were overly complex and only appeared to work correctly (because we had not formulated good tests): we generated the indented tree (with duplicate rows for concepts with multiple parents) at the server, hiding subtrees after a certain depth or where direct children of a given node exceeded a certain number. Through trial and error we found that limiting concept graph download to edge lists and concept metadata worked, if a little sluggishly, even for our largest value sets. From there, we were a little surprised to also discover, performing graph operations using graphology.js to generate indented rows and calculate descendant records for collapsed descendants was very fast.

We had long wondered why other value set browsing/editing tools didn’t directly display selected concepts in the context of their vocabulary hierarchies; it seems such an obvious and beneficial feature. The answer is, apparently, that it’s hard. Value sets range vastly in size and graph topology (width, depth, connectedness, polyhierarchy); designing a user interface that handles all cases well is a serious challenge. This has proven to be an historical development challenge, with few implementations to our knowledge. Nevertheless, we persevered to the point of proving that it can be done and have built an interface that does a credible job of it, with plenty of room for improvement of course.

As to why previous tools hadn’t used a tabular presentation to compare concept membership across multiple value sets, getting it to work required creative use of React, which is a more flexible UI framework than those used by other value set authoring tools, and this display makes the need for hierarchical concept presentation more painfully clear.

An application of this scale should not be built by one and a third programmers but by a software development team including project management; dev ops; quality assurance to implement unit, end-to-end, and user testing. A lot of mistakes and blind alleys could have been avoided, especially if we had been able to begin with a test-driven development approach. On the other hand, the application has provided significant value to our group and to the N3C community at large, and we built it with the resources we had.

Frontend caching was essential; sluggish initial load times for sizable value sets would be tolerated, but when every page reload or change in the list of selected value sets took just as long, users grew frustrated. An early approach used a library that cached each distinct API call (accounting for differences in parameters) but wasn’t sufficient. We built an overly complex but functional system for caching results granularly so that subsequent calls for overlapping data could use the cache and only download items that had not been retrieved. That is, if a related value set is added, it is only necessary to retrieve metadata for concepts not contained in the value sets already selected. We encountered many problems caused by cached data that should have been cleared for various reasons. We tried a variety of solutions including, horribly, asking users to hit an Empty cache button on the help page whenever the application misbehaved in case bad cache data was the culprit. Our current imperfect strategy is simply to automatically clear the user’s cache every 24 hours. Clearing least recently used cache data would have been helpful, but the complexity of our granular caching approach makes it difficult to implement.

### Limitations and future work

VS-Hub needs additional features for users to explore vocabularies for candidate concepts by string search or by exposing concepts related to those displayed but not currently included in the subgraph.

Hierarchical concept display makes it considerably easier to author parsimonious intensional value sets, which we consider a best practice, but it needs to go further and help users identify common parent or ancestor concepts when a more parsimonious definition is possible. The approaches we have tried so far have compromised usability by adding excessive polyhierarchy or bringing in unwanted descendants.

When vocabulary changes lead to changes in value set expansion, VS-Hub makes it easy to see these changes in context, but users want to understand, especially, why certain concepts no longer appear in the expansion and help finding replacements if appropriate. We have not yet tried to address this need.

The current implementation has high memory demands.

We envision VS-Hub being generalized and used more widely by 1) accommodating and storing data from more value set formats (VSAC, FHIR, etc.); 2) connecting it to external value set repositories; 3) allowing users to build and store value sets optionally synced to theN3C Enclave and other repositories; 4) allowing term usage counts and value set patient/record counts from multiple sources; and 5) by allowing institutions to host their own VS-Hub instances, connected to whatever private or public value set repositories, analysis platforms, and data sources they like. These extensions will make VS-Hub useful to a very wide community of RWD researchers and analysts. We do not currently have resources to implement them but hope to attract open-source software developers who could help.

VS-Hub represents a significant advance in the technology available for analyzing and authoring value sets. It removes obstacles that value set developers face in following best practices and making use of prior work. We have demonstrated the importance of the features it introduces, reviewed challenges encountered in implementing them, and provided lessons learned to ease the path of others who may attempt similar work. We invite open-source software developers to join us in bringing VS-Hub to a much wider community.

## Data Availability

All data produced in the present study are available upon reasonable request to the authors

## Acknowledgements

We thank the N3C Data Liaison and Palantir implementation teams for their feedback and help making VS-Hub possible. This work was supported by the National Institutes of Health (NIH) Agreement OT2HL161847 01 and NCATS U24 TR002306.

## Footnotes

1 Observational Health and Data Science (OHDSI),^1,2^ The National Patient-Centered Clinical Research Network (PCORNet),^3^ All of Us^4^, and The National COVID Cohort Collaborative (N3C)^5^

† We refer to defined sets of controlled medical vocabulary codes as value sets, though the N3C and OHDSI communities call them concept sets, the literature specifically discussing them in the context of RWD research generally calls them code sets, and some ontology communities call them enumerations. In other contexts they may also be called code lists, groupers, or term sets.

* The software has been called TermHub and that name lingers in various places; we changed it to VS-Hub to avoid possible trademark infringement.

* Ontology Access Kit is cited in the references.^14^ The other tools listed are easily found using any search engine.

* Though concepts have many types of relationships, VS-Hub currently displays only is-a/subsumes and other hierarchical relationships captured in the OMOP `concept_ancestor` table.

† We are also working on a node-link diagram that will more intuitively convey the DAG structure.

* We have this as a NetworkX directed graph generated from all rows of the `concept_ancestor table where min_levels_of_separation = 1.`

